# Enabling global-scale nucleic acid repositories through versatile, scalable biochemical selection from room-temperature archives

**DOI:** 10.1101/2024.04.12.24305660

**Authors:** Joseph D. Berleant, James L. Banal, Dhriti K. Rao, Mark Bathe

## Abstract

Conventional collection, preservation, and retrieval of nucleic acid specimens, particularly unstable RNA, require costly cold-chain infrastructure and rely on inefficient robotic sample handling, hindering downstream analyses. These generate critical bottlenecks for global pathogen surveillance and genomic biobanking efforts, prohibiting large-scale nucleic acid sample collection and analyses that are needed to empower pathogen tracing, as well as rare disease diagnostics^1^. Here, we introduce a scalable nucleic acid storage system that enables rapid and precise retrieval on pooled nucleic acid samples—stored at room-temperature with minimal physical footprint^2,3^—using versatile database-like queries on barcoded, encapsulated samples. Queries can incorporate numerical ranges, categorical filters, and combinations thereof, which is a significant advancement beyond previous demonstrations limited to single-sample retrieval or Boolean classifiers. We apply our system to a pool of ninety-six mock SARS-CoV-2 genomic samples identified with theoretical patient data including patient age, geographic location, and diagnostic state, allowing rapid, multiplexed nucleic acid sample retrieval in a scalable manner to empower genomic analyses. By avoiding expensive and cumbersome freezer storage and retrieval systems, our approach in principle scales to millions of samples without loss of fidelity or throughput, thereby supporting the development of large-scale pathogen and genomic repositories in under-resourced or isolated regions of the US and worldwide.

## INTRODUCTION

Large-scale collection, transport, storage, and retrieval of nucleic acids is essential to enable numerous advanced biotechnological application areas including population-scale disease tracking^4^, precision genomic medicine^5–7^, forensics^8^, and global ecological record-keeping^9,10^. In particular, genomic DNA and RNA collected from large and diverse patient cohorts empower both pathogen tracing and genomic medicine by enabling the prediction of disease-onset likelihood, as well as informing personalized treatment plans^7,11–15^. In general, intact native nucleic acid samples provide the most complete representation of genomic information, including epigenetic marks, such as methylation patterns in DNA and genomic viral RNA, as well as modifications to mRNA and lncRNA^16,17^.

Comprehensive nucleic acid analyses rely on both short-read and long-read sequencing platforms to generate the most detailed and accurate genomic data. These platforms, along with gold standard assays like mass spectrometry for analyzing epigenomic and RNA modifications^18^, require energy-intensive cold-chain infrastructure for sample preservation and transport to centralized analysis facilities^19–21^. This infrastructure burden is particularly challenging for RNA samples, which are highly susceptible to degradation without stringent preservation protocols^22,23^. Even after reaching analysis facilities, samples require prolonged low-temperature storage while awaiting comprehensive analyses, which are often both time- and cost-intensive. Further, the throughput and efficiency of sample recall from large-scale, automated freezer systems are limited by mechanical factors such as robotic automation speed.

As epidemiology, pathogen surveillance, personalized medicine, and ecological conservation efforts scale to worldwide sample collection, the aforementioned challenges create significant technological barriers that limit access to nucleic acid samples from under-resourced regions both in low- and high-income nations^24^. This contributes to severely limited participation in rare disease research, where understanding complex genetic traits and disease associations requires the analysis of tens of millions or more intact DNA and RNA samples from diverse global populations^25–29^. Similarly, pathogen monitoring and ecological preservation efforts at a global scale become prohibitively cumbersome and costly. Thus, there is an urgent need for low-cost, low-energy, and scalable storage infrastructures that preserve DNA and RNA at the point of collection while simultaneously enabling ambient transport and efficient sample retrieval for downstream genomic analyses.

Traditional biosample storage methods, which rely on barcoded tubes stored in freezers or liquid nitrogen tanks, face significant challenges in cost and practicality as collections scale into the millions. For these vast biosample databases, sophisticated systems are required for efficient sample search and retrieval, in addition to continuous energy consumption and high-cost storage infrastructure. While automation partially alleviates these barriers, these biosample databases remain limited by their low storage density and use of sequential rather than parallel sample access.

In contrast, molecular-based approaches enable the pooling of millions to billions of unique nucleic acid biosamples per tube, effectively creating a highly dense biosample database. Retrieval of specific samples or sets of related samples can be achieved using biochemical approaches such as PCR^30^, magnetic pulldown^31^, or fluorescence-activated sorting (FAS)^2,3^ which use molecular labels like primers, affinity tags, or fluorescent dyes. Primers and affinity tags are often made with DNA, capitalizing on the innate specificity of DNA hybridization to ensure precision and scalability of retrieval. These methods simultaneously process the entire pool of biosamples, significantly improving retrieval efficiency by executing millions to billions of concurrent molecular search and retrieval operations in solution, far surpassing the capabilities of conventional manual or robotic search and retrieval of individual tubes. For example, we have previously demonstrated a system of silica-encapsulated biosamples labeled with DNA barcodes. The DNA barcodes encode metadata about each sample, facilitating precise identification with fluorescent probes, while silica encapsulation effectively preserves biosamples at room temperature and prevents unintended interactions with the biosamples that could interfere with biochemical retrieval. With this framework, we demonstrated Boolean retrieval in two studies: one using a prototypical image database stored in plasmid DNA^2^, and another with a pool of genomic DNA and RNA samples^3^. Retrieval was demonstrated to have high specificity, with successful retrieval of target samples comprising just 1 in 10^6^ of the total sample pool^2^. Despite their advantages, this and related molecular labeling systems^30,32,33^ remain severely limited due to their inability to perform arbitrary, complex queries such as routinely performed on modern digital databases using Structured Query Language (SQL). Such complex retrieval operations are essential for genomics and epidemiological applications in which multiple diagnostic conditions and/or time periods are sought from very large-scale genomic databases that reach tens to hundreds of millions or even billions of samples or patients.

To enable biosample database implementations that approach the capabilities of modern digital databases, here we develop a scalable biosample database that permits metadata queries for numerical ranges, such as ranges of dates or ages; categories, such as cities or countries; and our previously demonstrated Boolean classifications, such symptomatic/asymptomatic. Using a synthesized database of model SARS-CoV-2 genomes, we demonstrate the expressiveness of our query system in a simulated pathogen outbreak scenario, where passengers entering a major international airport are comprehensively swabbed for SARS-CoV-2 to track the pandemic (**Figure 1a**). Each sample simultaneously encodes age, vaccination status, presence of symptoms, and flight information like flight number, date, time, and place of origin. We performed retrospective epidemiological and immunological analyses with queries of increasing complexity, including queries for a particular health status, for three distinct age ranges, and for simultaneous matches to criteria for date range, location, and health status. These examples illustrate a general-purpose SQL-like query language, permitting arbitrary logical expressions composed of numerical range, categorical, and Boolean metadata criteria. This work thereby demonstrates a scalable storage system that supports the expressive query capabilities of modern digital databases (**Figure 1b**), while eliminating cold-chain logistics through silica encapsulation for long-term RNA sample preservation at room temperature^2,34^. This framework addresses bottlenecks in nucleic acid storage and retrieval, offering broad applications in molecular diagnostics, pathogen surveillance, and ecological preservation.

**Figure 1.**
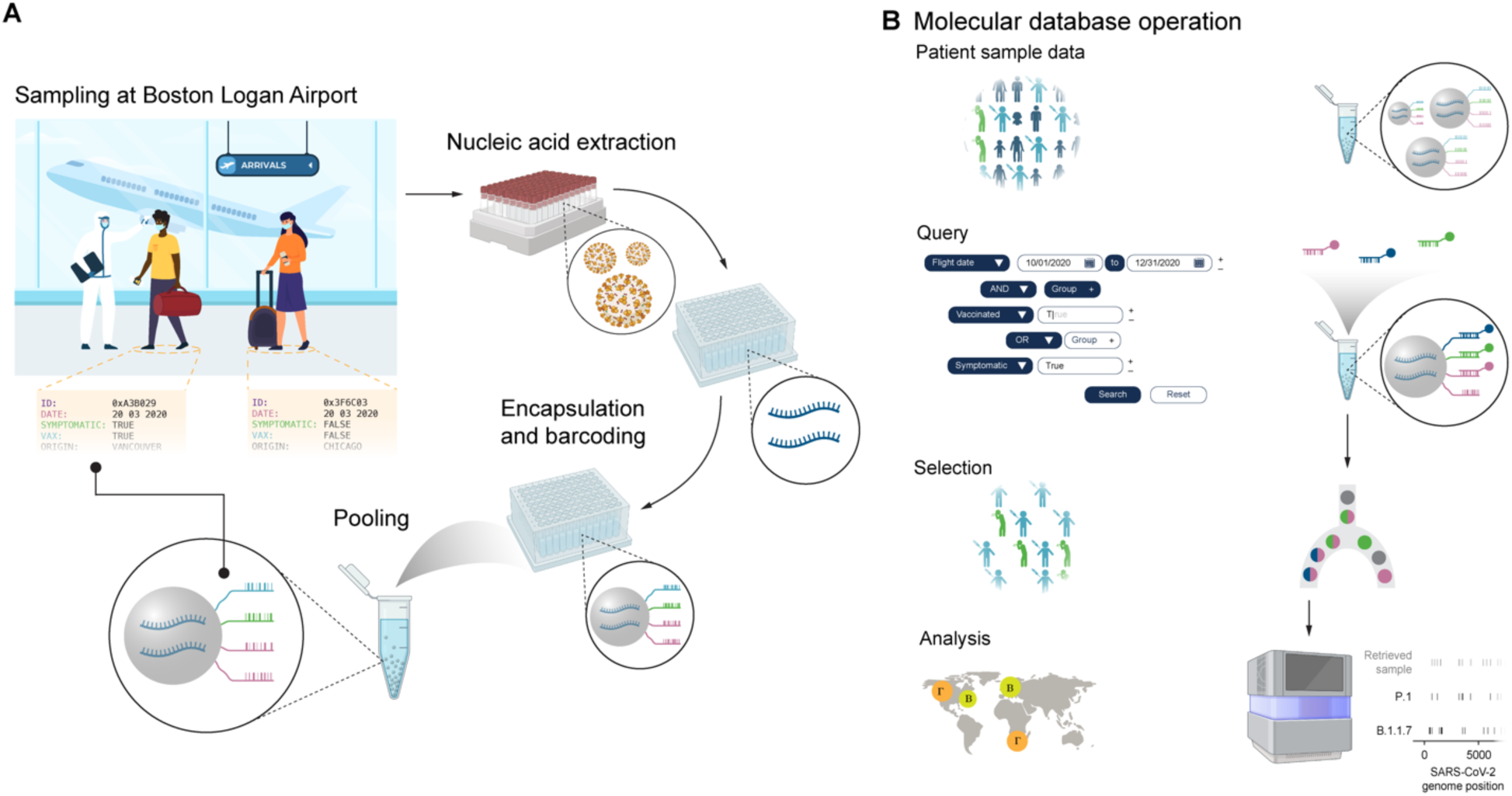
| Application of a molecular database to simulated SARS-CoV-2 tracking. **a**, Simulated scenario of sample collection at Boston Logan airport with subsequent pooling and nucleic acid extraction, encapsulation, and barcoding of samples using our proposed molecular filesystem. **b**, Workflow for querying and analyzing samples within a molecular database, shown side-by-side with generic database operations.

### Database construction

Ninety-six mock “patient samples” were separately encapsulated in 5 µm silica microcapsules (**Figure 1a**). These microcapsules simulated samples drawn from passengers flying into Logan International Airport in Boston, Massachusetts, and subsequently stored archivally for future diagnostic or epidemiological testing, if needed. Each sample consisted of at most one variant of the SARS-CoV-2 genome using either synthetic P.1 and B.1.1.7 variants and a unique internal 85-nt barcode whose purpose was to aid validation of microcapsule retrieval by making possible downstream identification of de-encapsulated microcapsules in a pool. Variants were quasi-randomly assigned to each sample, with approximately a 15% chance of some variant being present. Encapsulation followed the procedure previously used (see Methods)^2,34^. In contrast to our previously encapsulated nucleic acids^2,34^, the DNA encapsulated here consisted of linear DNA of differing lengths: short internal barcodes of 85-nt that represented unique identification barcodes for each “patient” and 5-kb fragments of long genomic synthetic SARS-CoV-2 RNA. However, because the encapsulation process relied only on charge interactions between the negatively charged phosphate backbone of nucleic acids, we expected successful incorporation of all nucleic acids irrespective of length (**Supplementary Figure 1**), as previously demonstrated^34^. We validated nucleic acid encapsulation and de-encapsulation with approximately 1 copy of SARS-CoV-2 genome and ∼10^8^ copies of the internal barcode per microcapsule.

### Database labeling and querying

The ideal SARS-CoV-2 genomic database would label each sample with metadata features, such as its unique identifier, patient health status, sample acquisition date, and flight origin. For our example use case, several features were chosen to describe a variety of relevant metadata, including patient age and month and year of the arrival flight, which are examples of numerical metadata; vaccination status, which is an example of Boolean metadata; and flight number and city of origin, which are examples of categorical metadata. For each microcapsule, each feature value was encoded into a set of barcodes to be displayed on its exterior (**Figure 1a**), with a total of either 13 or 14 unique 25-nt ssDNA barcodes per microcapsule (see Methods). Differing encoding strategies were employed based on the type of metadata (numerical, Boolean, or categorical), which enabled type-specific queries of each feature, such as queries for numerical ranges. More detail about each encoding and retrieval strategy is given in Methods.

For numerical metadata, we encoded each numerical value as a sequence of digits in a mixed-radix representation, corresponding to a sequence of barcodes with one barcode per digit. Queries for an exact numerical match are performed by probing for every barcode in its representation, while ranges of various sizes are possible by omitting one or more of the less significant digits (**Figure 2**). Categorical metadata were encoded using distinct combinations of *k* barcodes for each possible value, so that the value of *k* determined the number of barcodes required to identify a sample’s specific feature value. This approach scales to accommodate an extensive number of feature values. Boolean metadata, on the other hand, were straightforward, with the presence or absence of a single barcode indicating the feature’s state.

**Figure 2.**
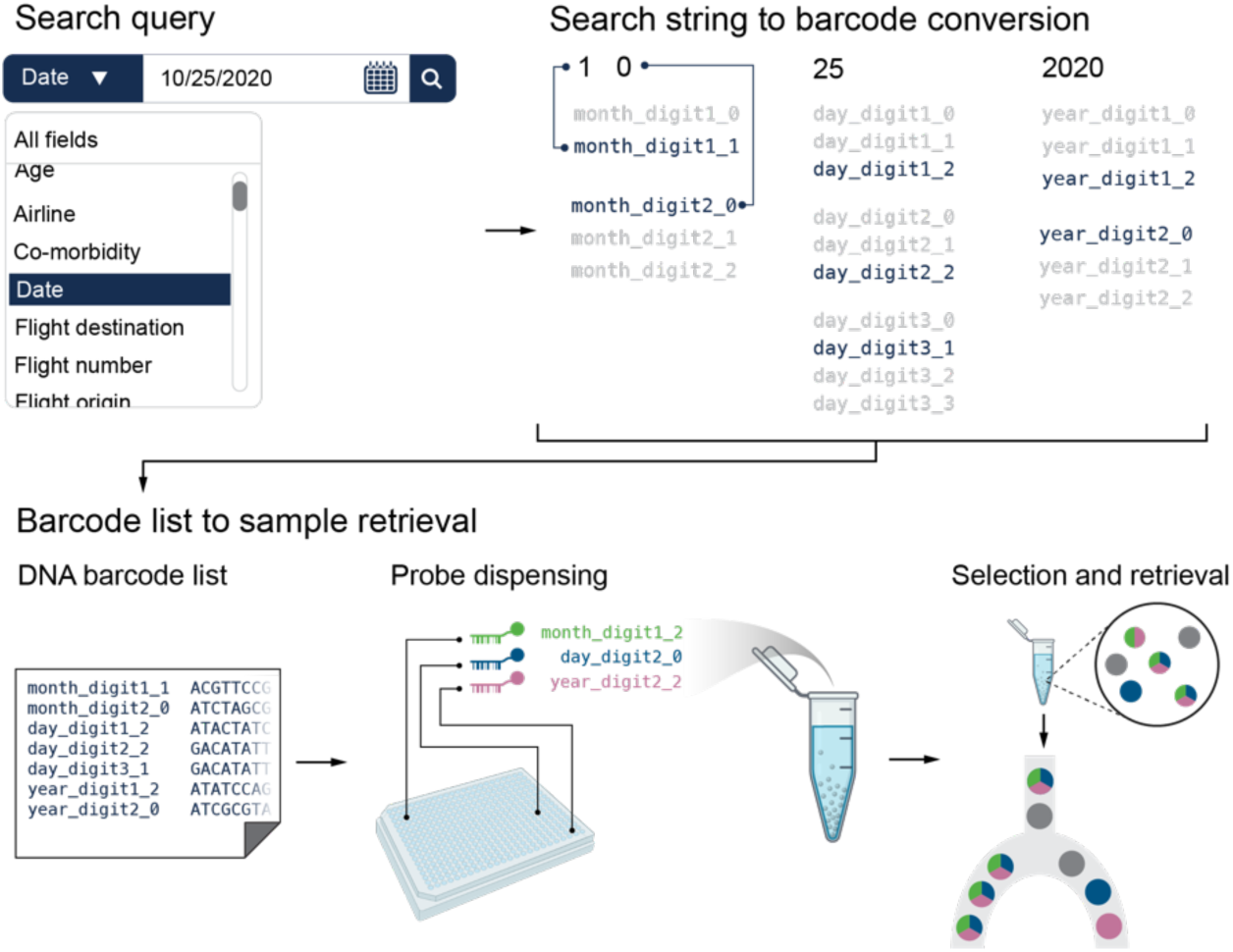
| Conversion of search query to operations on a molecular database. Numerical, Boolean, and categorical metadata are converted into a corresponding logical expression comprising AND, OR, NOT operations on several barcodes. This logical expression then guides the dye-labeling strategy, search grouping, and dispensing of dye-labeled DNA barcodes for sample selection. Selected samples are then retrieved using optical sorting. In this schematic, a specific date is represented by a total of seven barcodes (two for the month, three for the day, and two for the last two digits of the year). Here, month and year are represented in base 10 while the day is represented in a combination of bases 3 and 4. Ranges of contiguous dates are specified by omitting one or more barcodes.

When performing a query, the search string is first translated into a DNA barcode list (**Figure 2**). This list not only facilitates the query but also informs the selection of unique dyes needed for FAS. For NOT logic, the barcode that denotes the unwanted condition is tagged with a fluorescent dye. During sorting, microcapsules that do not show this fluorescence are selected, thus excluding the condition indicated by the dye. OR logic employs a single dye for all relevant barcodes, selecting samples with the matching dye. For AND logic, each query barcode is tagged with a distinct dye, and only samples displaying all unique dyes are selected. This intricate logic necessitates a careful selection of dyes and strategic grouping of search parameters, ensuring precise sample retrieval.

### Immunological case study

To demonstrate the application of a large-scale molecular database with advanced search query capabilities, we performed several database queries on the synthetic database of 96 SARS-CoV-2 samples hypothetically collected from airplane passengers entering Boston, MA. We designed search queries to demonstrate the breadth of the queries enabled by the sample labeling approach, as well as to show how an actual database of this type could be used to answer valuable retrospective epidemiological and immunological questions.

We began with the immunological question of whether specific SARS-CoV-2 variants were present in asymptomatic passengers. To answer this question, we used a query for when the Boolean feature “symptomatic” was false. For each sample, the barcode “bc_symptomatic” was present when this feature was true, and absent when this feature was false. Thus, our query “NOT symptomatic” should retrieve exactly those samples not displaying this barcode.

The presence of the barcode “bc_symptomatic” on each microcapsule was determined using a fluorescent probe combined with fluorescence-associated sorting (FAS). However, in comparison to previous work using FAS for barcode detection on microcapsules^2,34^, the number of distinct barcodes attached per microcapsule was increased from 3 to up to 14, leading to a proportional decrease in the copy number of each barcode per microcapsule. To compensate for this copy number reduction, we used fluorescence amplifiers routinely used to detect low copy numbers of RNA in cells in flow cytometry^35^ (**Figure 3a**). The pool of microcapsules was mixed with this amplifier probe and subsequently passed through a FAS instrument, which generates a stream of droplets. Fluorescence from each droplet was measured, and selected populations were defined using fluorescence intensity to distinguish microcapsules with and without the fluorescent probe attached. The low-intensity population was separated from the rest of the pool for subsequent de-encapsulation and sequencing.

**Figure 3.**
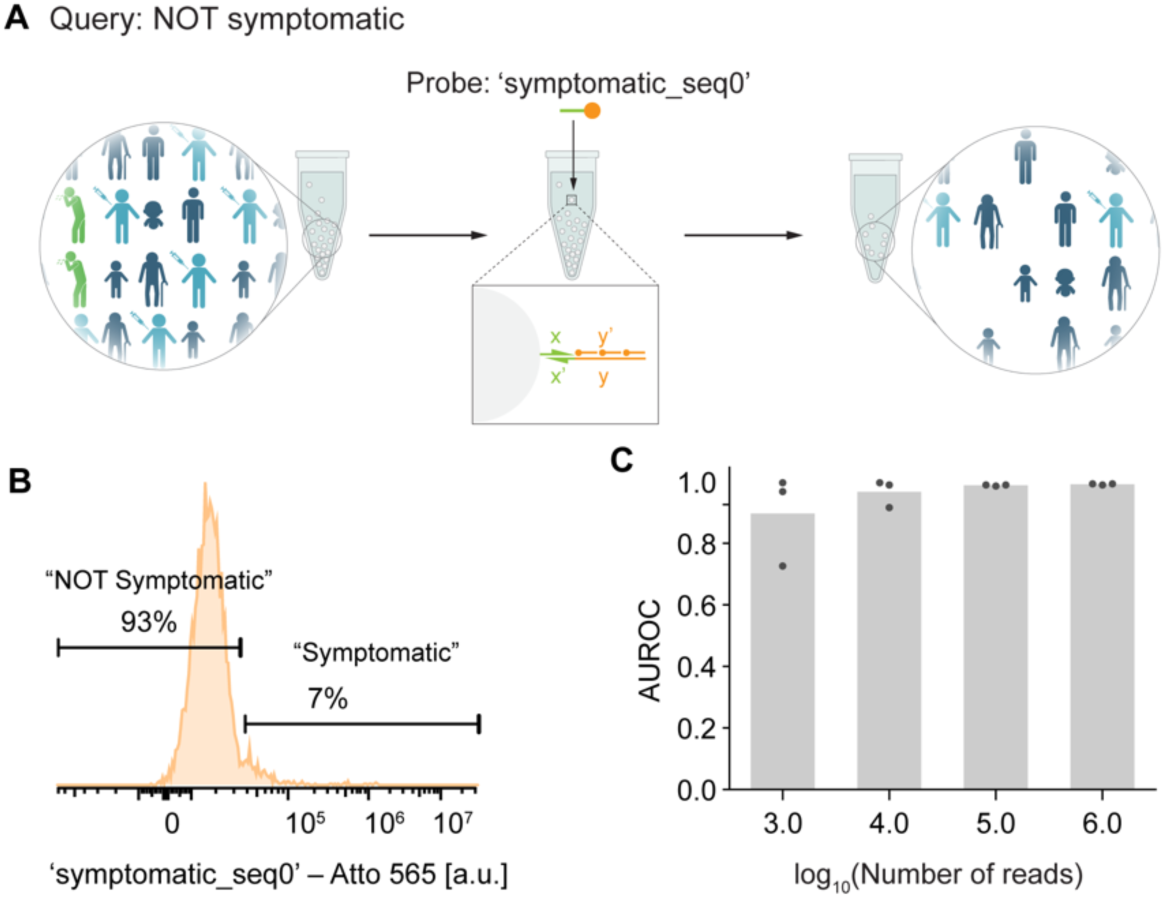
| Database querying results for “NOT symptomatic”. **a**, For this query, the probe included a region complementary to barcode “bc_symptomatic” followed by a repeating sequence that allowed 3-fold fluorescence amplification. **b**, Histogram of FAS results showing 93% of microcapsules with low fluorescence indicating absence of the “bc_symptomatic” barcode. **c**, The area under the ROC curve (AUC), which measures quality of a binary classifier, increases as more sequencing reads are processed. This is expected behavior as the variance in the number of reads for each sample is reduced with more reads.

To validate correct retrieval, sequencing reads of the 85-nt internal barcodes were matched to the known sequences to identify the samples in the sequencing data. The number of reads of each sample showed clear separation between the retrieved and non-retrieved microcapsules (**Figure 3b**). To quantify the retrieval performance, receiver operator characteristic (ROC) analysis of the number of reads per sample was performed and the area under the ROC curve (AUC) computed. The AUC value is interpreted as a quality score between 0.0 and 1.0, with 1.0 indicating ideal behavior (all correct samples were present at higher levels than any incorrect samples) and 0.0 indicating maximally incorrect behavior (all incorrect samples present at higher levels than correct samples) (**Figure 3c**). For this query, the mean AUC over 3 replicates was computed as increasing numbers of sequencing reads were processed. The AUC was high (AUC=0.99) when the number of sequencing reads surpassed 10^5^, indicating enrichment of the desired populations.

Next, we demonstrated how range queries on patient age could be used to explore if certain age groups were more susceptible to different SARS-CoV-2 variants^36^. We considered three age range queries of different size: “age = 0”, “15 ≤ age ≤ 19”, and “50 ≤ age ≤ 74”. For the narrowest query “age = 0”, we selected samples labeled with barcodes bc_age_x25_seq0, bc_age_x5_seq0, and bc_age_x1_seq0 using the same type of multi-stranded branched probes as previously described, labeled with fluorophores Atto 565, Alexa Fluor 647, and Alexa Fluor 750, respectively. For the moderate-range query “15 ≤ age ≤ 19”, we selected samples labeled with barcodes bc_age_x25_seq0 and bc_age_x5_seq3, using probes labeled with fluorophores Atto 565 and Alexa 647. For the query of broad range “50 ≤ age ≤ 74”, we selected samples labeled with the barcode bc_age_x25_seq2 using a probe labeled with fluorophore Atto 565. In each case, sequencing the internal 85-nt barcodes showed that correct samples were retrieved, as indicated by AUC values of 1.0, 1.0, and 0.9, respectively, for at least 10^5^ sequencing reads (**Figure 4**).

**Figure 4.**
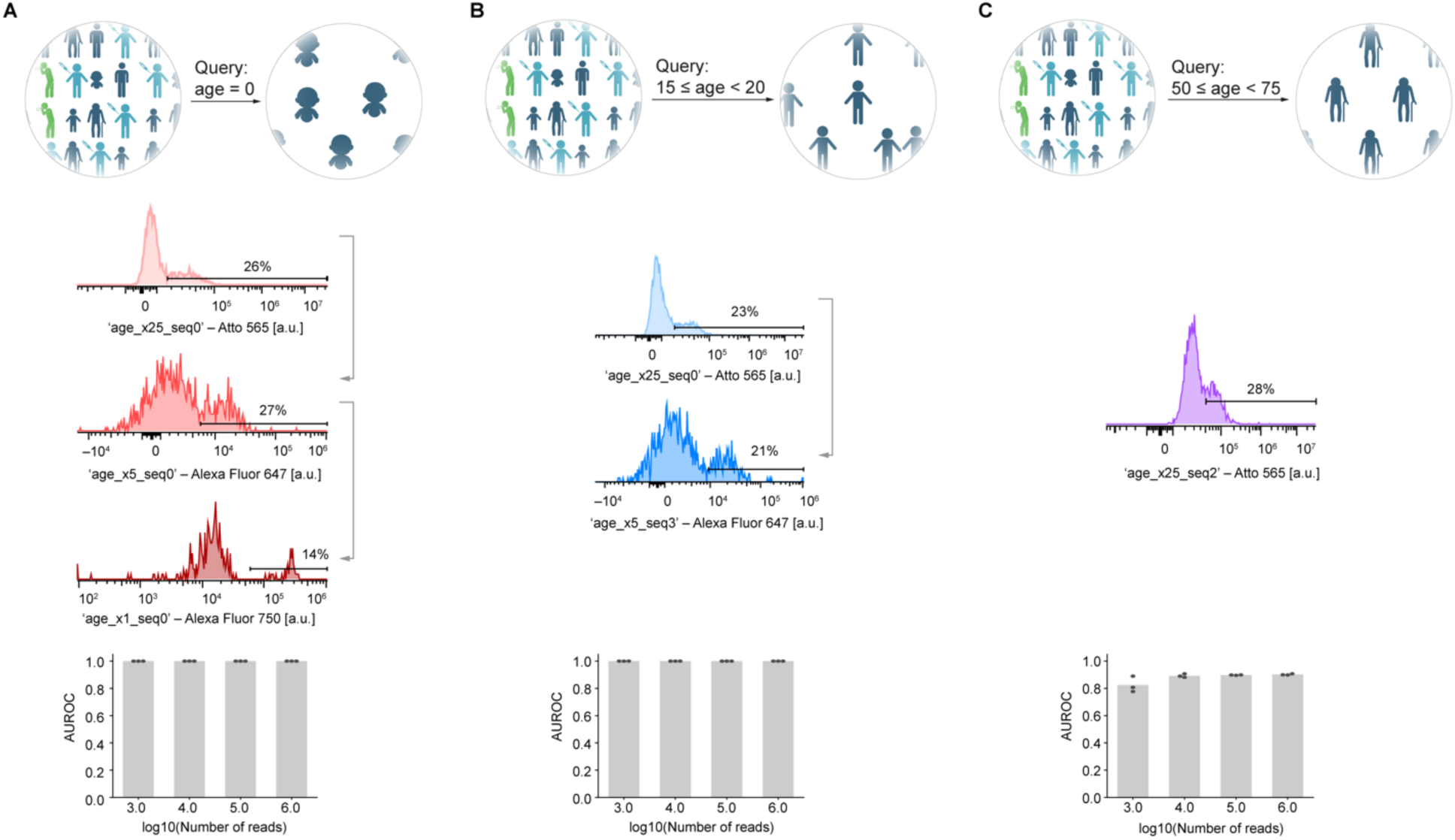
| Age range queries, demonstrating range queries of different size. **a**, “age = 0”, **b**, “15 ≤ age ≤ 19”, and **c**, “50 ≤ age ≤ 74”.

### Epidemiological case study

We then sought to demonstrate how a composite query involving multiple metadata types can be used to understand the transmission of SARS-CoV-2 infection from specific areas, date ranges, and flight city^37^. For the epidemiological case study, we illustrated two aspects of our database: first, efficient representation and querying of categorical features; and second, the composition of several smaller queries into arbitrarily complex logical expressions. This was performed via retrieval of all samples for passengers flying from Chicago between July and September 2020, who were either symptomatic or unvaccinated. This was equivalent to the query “(symptomatic OR NOT vaccinated) AND flight_city = Chicago AND 6 ≤ arrival_month ≤ 8 AND arrival_year = 2020”, which combines queries on two numerical features, one categorical feature, and two Boolean features. Such a query necessitated examining eight barcodes: bc_vaccinated, bc_symptomatic, bc_city_seq0, bc_city_seq3, bc_city_seq4, bc_flight_month_x3_seq2, bc_flight_year_x10_seq2, and bc_flight_year_seq0, making it the most complex query tested on any molecular database to the best of our knowledge, both semantically and in terms of the number of barcodes tested. To further exhibit the flexibility of our approach to a variety of fluorescent channels and probe design methodologies, we selected a new set of dyes for this query, which modified the bandwidth of our dye markers (**Figure 5a**). Specifically, we transitioned from Atto 565, Alexa Fluor 647, and Alexa Fluor 750 to Atto 488, Atto 565, and Alexa Fluor 647. The reduced brightness of Atto 488 relative to the other dyes necessitated the use of branched probe designs that amplify fluorescence signals, similar to branched probed designs used to improve the relative brightness of low-copy targets in cell imaging^38,39^. This strategy allowed the amplification of the net fluorescent signal by increasing the number of dye markers per barcode.

**Figure 5.**
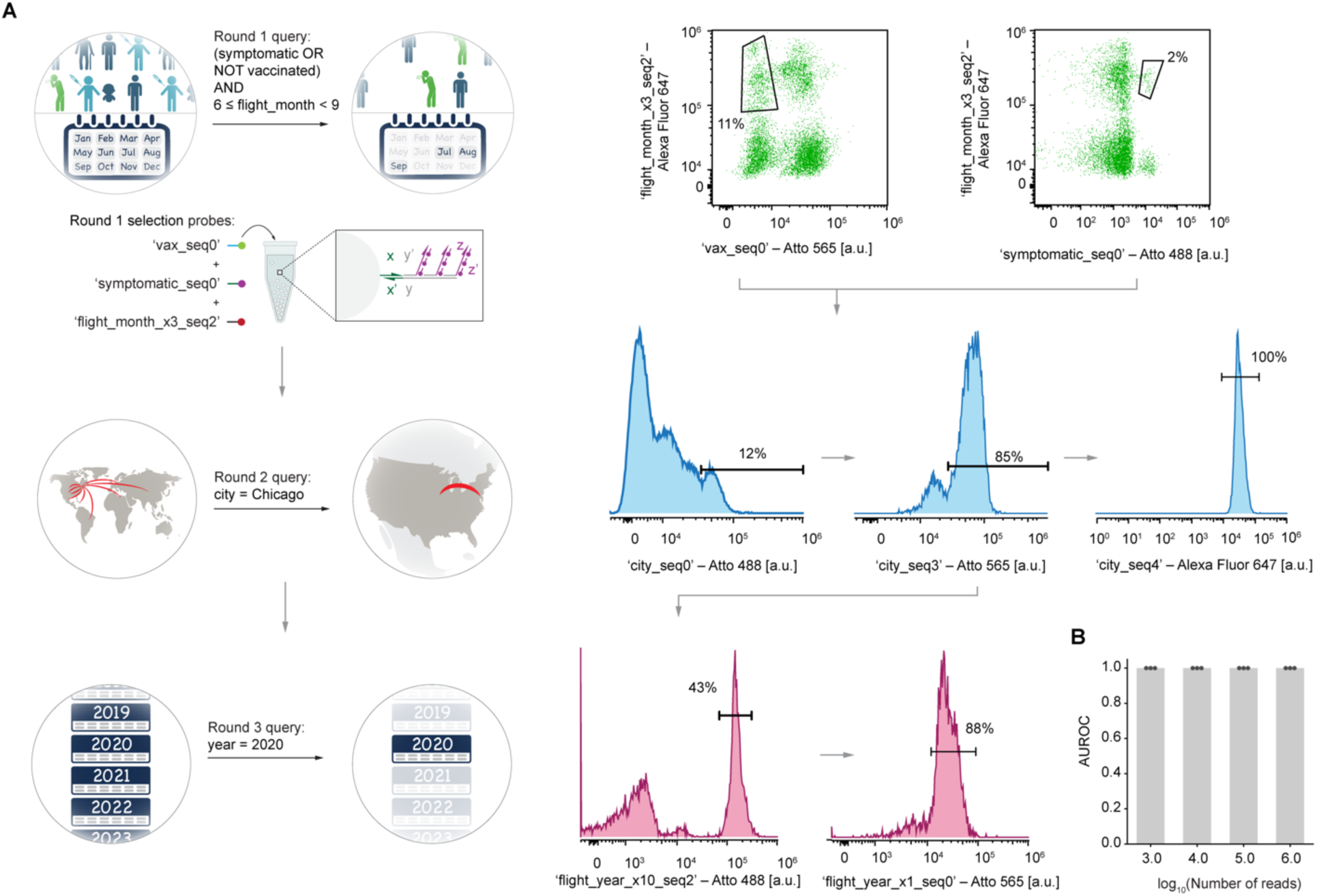
“(symptomatic OR NOT vaccinated) AND flight_city = Chicago AND 6 ≤ arrival_month ≤ 8 AND arrival_year = 2020”. **a**, For this query, branched probes providing 9-fold fluorescence amplification were used. Scatterplots (top-right) from the first round of FAS illustrate the cluster separation observed. Histograms for the second and third rounds of FAS are shown (middle, bottom). **b**, The AUROC score remains at 1.0 down to 1000 reads, indicating all correct samples had higher sequencing counts than all incorrect samples.

This query was performed over three FAS passes, using probes with fluorophores Atto 488, Atto 565, and Alexa Fluor 647 for bc_vaccinated, bc_symptomatic, and bc_flight_month_x3_seq2, respectively, for the first pass. The sorted populations were stripped of their fluorescent probes using a denaturation buffer. For the second round of selection, fluorophores Atto 488, Atto 565, and Alexa Fluor 647 were used for selecting bc_city_seq0, bc_city_seq3, bc_city_seq4, respectively. Again, the previous probes were removed prior to subjecting the sorted population to the next round of selection. For the final FAS pass, Atto 488 and Atto 565 were used for bc_flight_year_x10_seq2 and bc_flight_year_seq0, respectively. Sequencing of the 85-nt internal barcodes after all three passes indicated correct retrieval of all samples (AUC = 1.0) (**Figure 5b**). This demonstration illustrates two important features of our approach: first, the ability to implement a single molecular database query that describes criteria spanning many features, such as a numerical feature falling in a specified range and multiple Boolean features having the desired truth values; and second, the ability to split complex queries over several FAS passes without loss of retrieval fidelity.

### SARS-CoV-2 sequencing

Identifying the dominant SARS-CoV-2 variant is essential for assessing the virulence of emerging strains, forecasting outbreaks, and expediting vaccine development^40^. We aimed to identify the predominant SARS-CoV-2 variant in our queries, given that our samples were encapsulated with either the Alpha or Gamma variants. After sorting 100,000 to 700,000 microcapsules for each selection, we selectively sequenced samples that yielded a positive result from a specifically designed end-point tiling amplicon PCR for SARS-CoV-2^41^ (**Supplementary Figure 3**). Computational demultiplexing using Freyja^42^ revealed that all samples showed mostly Alpha variants followed by Gamma variants and other variants that were not assigned by Freyja (**Figure 6a**). In all cases, the ratio of expected Alpha to Gamma abundance: 8:3 for selections from **Figure 3**, 2:1 for selections from **Figure 4c**, and 1:0 for selections from **Figure 5**, closely matched the measured abundances in **Figure 6a**, further providing support to the high retrieval precision observed using an orthogonal sequencing approach. Identifying the dominant SARS-CoV-2 variant is essential for assessing the virulence of emerging strains, forecasting outbreaks, and expediting vaccine development^40^. We aimed to identify the predominant SARS-CoV-2 variant in our queries, given that our samples were encapsulated with either the Alpha or Gamma variants. After sorting 100,000 to 700,000 microcapsules for each selection, we selectively sequenced samples that yielded a positive result from a specifically designed end-point tiling amplicon PCR for SARS-CoV-2^41^ (**Supplementary Figure 3**). Computational demultiplexing using Freyja^42^ revealed that all samples showed mostly Alpha variants followed by Gamma variants and other variants that were not assigned by Freyja (**Figure 6a**). In all cases, the ratio of expected Alpha to Gamma abundance: 8:3 for selections from **Figure 3**, 2:1 for selections from **Figure 4c**, and 1:0 for selections from **Figure 5**, closely matched the measured abundances in **Figure 6a**, further providing support to the high retrieval precision observed using an orthogonal sequencing approach.

**Fig. 6.**
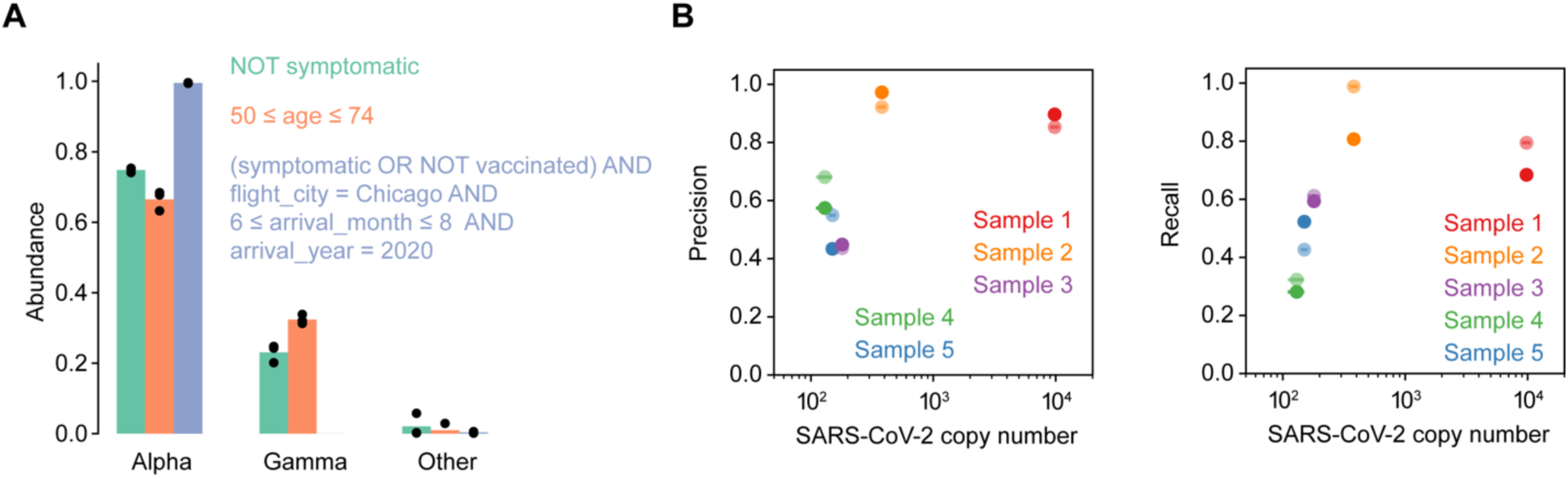
Sequencing results for synthetic and clinically-derived SARS-CoV-2 samples that were encapsulated and then de-encapsulated, to demonstrate the feasibility of our approach to real-world nucleic acid samples. (**A**) For the three database queries “NOT symptomatic”, “50 ≤ age ≤ 74”, and “(symptomatic OR NOT vaccinated) AND flight_city = Chicago AND 6 ≤ arrival_month ≤ 8 AND arrival_year = 2020”, the variants in each sample were quantified. These variants should correspond to the synthetic Alpha or Gamma that had been encapsulated in each of the 96 mock patient samples. (**B**) Results for sequencing of clinical SARS-CoV-2 samples that had been encapsulated and then de-encapsulated. Dark and light-colored circles represent each duplicate sequencing run for each sample.

To demonstrate the viability of encapsulation for storage and recovery of clinical SARS-CoV-2 samples, we applied our approach to five distinct patient-derived samples. Each sample contained different sub-lineages of the Omicron variant, allowing us to gauge our ability to detect small variations in the viral genomes present during encapsulation through the comparison of called variants of each sample with and without encapsulation. For each sample, we successfully recalled the sub-lineages of Omicron variants (**Supplementary Figure 2**). Further examination of the data indicated that the precision and recall of variant calling for the encapsulated samples were affected by the sequencing coverage (**Figure 6a**, top and **Figure 6b**, bottom), which we attribute to the low copy numbers of SARS-CoV-2 retrieved from encapsulation resulting in amplicon dropouts (**Supplementary Figure 4** and **Supplementary Figure 5**).

## DISCUSSION AND CONCLUSION

The large-scale, global collection of genomic DNA and RNA across health and security sectors ranging from pathogen surveillance to personalized medicine now calls for scalable, low-cost long-term storage and efficient sample retrieval capabilities. In this work, we demonstrated an intuitive yet powerful sample labeling strategy that significantly expands querying capabilities within a pooled molecular database, rendering it analogous to searching common digital file databases such as public datasets hosted by Google BigQuery, Microsoft Azure, and Amazon Web Services^43–45^. The nucleic acid database query language can accommodate arbitrary logical combinations of ranged queries, categorical queries, and truth queries on features that have been encoded into barcodes displayed on each nucleic acid specimen, encapsulated within a microcapsule for long-term stability. In our demonstration of the search query language, we showed how this nucleic acid database could be applied to answer several examples of retrospective epidemiological and immunological questions by analyzing sequencing results from cohorts retrieved from a database of simulated SARS-CoV-2 samples.

Our demonstration used a database of 96 samples with up to 14 barcodes per microcapsule, which allowed all 96 samples to be consolidated into a single tube resulting in a hundredfold reduction in storage space while maintaining a broad range of sophisticated search capabilities, including combinations of numerical range, categorical, and Boolean queries. This scalability and efficiency in sample management have direct implications for enhancing biosurveillance strategies as proposed by the Nucleic Acid Observatory^46^. By enabling the pooling of thousands or millions of samples into fewer tubes, our approach offers a significant footprint reduction over methods that require a separate vessel for each sample, thereby streamlining the process of monitoring and responding to pathogenic threats. Consider the traditional storage of 1 million nucleic acid samples. Storing these samples in –80°C freezers, each accommodating 40,000 samples, would require 25 freezers. In contrast, the microcapsule architecture, capable of retrieval from a pool of 10,000 distinct nucleic acids in just one tube at room temperature as we have previously shown^2,34^, would only require 100 tubes for storage. An entire tube can be queried in around 15 minutes, assuming a FAS rate of 1000 microcapsules per second and a redundancy of 100 microcapsules per sample. This streamlined approach not only offers a potential solution to the challenges outlined by the Nucleic Acid Observatory in deploying biosurveillance approaches but also underscores the potential for our technology to facilitate rapid and efficient global health responses. The approach represents a significant advance in expanding our ability to store, organize, and access nucleic acids, implementing the ability to perform the search functions that are essential for modern digital databases. Moreover, with the ongoing transformation of immunoassays^47^ and spatial tissue data^48^ into DNA molecules, we anticipate that the encapsulation and barcoding approach that we have demonstrated here can be used to store and query a comprehensive range of genomic, transcriptomic, and proteomic data. Finally, the prospect of encoding digital data, such as health records, into DNA, envisages a future where vast biological information could be efficiently stored, marking a significant leap forward in the compact and versatile storage of biological information in the palm of the hand.

## METHODS

### General materials

All DNA oligonucleotides (oligos)—including internal barcodes, splint adapters, 5’-amino-modified DNA barcodes, dye-labeled probes, preamplifier probes, amplifier sequences, master forward and reverse primers (**Supplementary Table 1**), random hexamers, and 20-mer oligodeoxythymidine—were synthesized and processed by Integrated DNA Technologies (IDT). Specifically, 5’-amino-modified barcodes in Echo 384 polypropylene microplates and internal barcodes in 96-deep well plates were purchased as desalted and delivered in nuclease-free water at 1000 μM and in 1× TE at 500 μM, respectively. Dye-labeled probes, adapter sequences, and branch sequences were received in 1× TE at concentrations of 100 μM, 33 μM, and 10 μM, respectively. The master primers were desalted and resuspended at 100 μM with nuclease-free water, while random hexamers and 20-mer oligodeoxythymidine were purified using ion-exchange high-performance liquid chromatography and resuspended at 50 μM with nuclease-free water. SARS-CoV-2 RNA controls were sourced from Twist Bioscience with catalog numbers 103909 and 104044. All oligos were stored at –20°C. Silica particles of 5 μm diameter with hydroxy-terminated surfaces (catalog number: DNG-B017) were obtained from Creative Diagnostics, and *N*-[3-(Trimethoxysilyl)propyl]-*N*,*N*,*N*-trimethylammonium chloride (TMAPS; 50% methanol; catalog number: H66414) was acquired from Alfa Aesar. Chemicals such as tetraethyl orthosilicate (TEOS), *N*-(2-aminoethyl)-3-aminopropyltrimethoxysilane (AEAPTS), N-methyl-2-pyrrolidone (NMP), isopropanol, and ethanol were sourced from Millipore Sigma, bearing catalog numbers: 131903, 8.19172, 270458, 278475, and 459836, respectively. DBCO-PEG5-tetra ester (1260-10) and azidoacetic acid *N*-hydroxysuccinimide ester (1070-100) were from Click Chemistry Tools. Additionally, carbonate buffer (500 mM, pH 9.0; catalog number: J63899.AK) and saline sodium citrate (SSC, 20×; catalog number: 15557044) were purchased from Thermo Fisher, while dextran sulfate (50% in water; catalog number: S4030) came from Millipore Sigma, 10% Tween 20 from VWR (catalog number: 97063-980), formamide from VWR (catalog number: 97062-006), and 5M sodium chloride from VWR (catalog number: 97062-858). The preparation of ammonium-functionalized silica particles followed previously published procedures using TMAPS^2,34^.

### Internal barcode generation

A subset of primers, 700 in total, from the validated 240,000 primer library^49^ was checked for alignment against SARS-CoV-2 using BLAST^50^. The alignment XML output file from BLAST was then parsed to create a list of primers orthogonal to the SARS-CoV-2 genome. The resulting primer list was further filtered by selecting primers with 60–65°C melting temperature using the melting temperature module in Biopython.

Internal barcodes were generated by first picking a master primer pair from the filtered primer list, forward: 5’–GGCTATGAGACTGTTCGCTAATCAC–3’ and reverse: 5’– CCCTTTGTGGGCACAGTTTAGTCTC–3’, which flanked a unique barcode taken from the primer list. Five nucleotide randomer spacers (N) were also added between the master primers and the unique barcode to increase the sequence diversity of the internal barcodes for downstream sequencing. Together, the 85-nucleotide internal barcode sequence structure is GGCTATGAGACTGTTCGCTAATCACNNNNNUUUUUUUUUUUUUUUUUUUUUUUUUNNNNNCCCTTTGTGGGCACAGTTTAGTCTC, where U is the unique barcode sequence.

### 96-well encapsulation

For each well in a Nunc 96 deep-well polypropylene plates (Thermo Fisher; catalog number: 278752), a total of 2 mg of ammonium-functionalized silica particles and 800 μl of 0.1% (v/v) Tween 20 in nuclease-free water were added. A total of 5 nanomoles of each internal barcode were added to their respective wells. Ten million copies of SARS-CoV-2 standards were added to each randomly selected well. A volume of 10 μl of TMAPS and 5 μl of TEOS were then added to each well. The plate was then covered with a chemically resistant silicone mat (Axygen; catalog number: AM-2ML-RD-S) and shaken for four days at 1500 rpm using a BioShake iQ thermal mixer (Bulldog Bio; catalog number: 1808-0506).

### Encoding of features with DNA barcodes

A full list of features is given in **Supplementary Table 4.3**. For each feature, a set of barcodes was allocated from which a subset was drawn to encode each feature value. The sets of barcodes for each feature were disjoint from each other. The encoding strategy for feature values differed based on the type of the metadata (numerical, Boolean, or categorical), which enabled type-specific queries of each feature, such as queries for feature values matching particular numerical ranges.

#### Numerical features

Numerical features were encoded using a mixed-radix number system (i.e. a sequence of digits with the base allowed to vary between positions) (**Supplementary Figure 6a**). A position with base *n* was allocated *n* distinct barcode sequences, one for each of the *n* possible digit values at that position. Thus, a feature value represented by *k* digits was encoded on each microcapsule with a collection of *k* distinct barcodes. Ranges of varying size could be specified by allowing some number of the least significant digits to vary (i.e. placing “wildcards” at these digits) (**Supplementary Figure 6b**). Experimentally, any numerical range specified in this manner can be retrieved by omitting the corresponding complementary probes during sorting. The base at each position was chosen to provide a good compromise between compression (number of barcodes required on each microcapsule) and the variety of the range sizes that could be represented.

#### Boolean features

Each Boolean feature was encoded using a single barcode assigned to that feature, similar to barcoding previously used to indicate image content in a database of images^2^. When the value of a Boolean feature was TRUE for a sample, the barcode was displayed on the microcapsule; a value of FALSE was indicated by the absence of that barcode.

#### Categorical features

For each categorical feature we used a combinatorial number system^51^ to associate each possible feature value with a distinct *k*-combination drawn from a chosen set of *n* barcodes, where *k* is the number of distinct barcodes used to represent this feature on each microcapsule. The number of possible feature values that may be represented is (*^n^_k_*), which grows rapidly with both *n* and *k*. The combinatorial number system provides a method to associate each possible *k*-combination with a unique integer value between 0 and (*^n^_k_*) − 1. Each feature value was assigned a unique numerical index in that range, from which the corresponding *k*-combination of barcodes was determined. The values of *k* and *n* were chosen to provide a reasonable compromise between feature width (the number of barcodes required on a microcapsule to represent its feature value) and the number of barcodes that needed to be allocated for this feature.

### Barcoding of individual samples and pooling

After encapsulation, the plate was centrifuged for 1 minute at 1000 relative centrifugal force (rcf). The supernatant was removed then backfilled with 1000 μl of 0.1% (v/v) Tween 20. A volume of 10 μl of AEAPTS was then added. The plate was then covered with a chemically resistant silicone mat and shaken for 1 day at 1500 rpm using a BioShake iQ thermal mixer.

To wash the encapsulated microparticles after amino modification, the plate was centrifuged for 1 minute at 1000 rcf. The supernatant was removed and then backfilled with 1000 μl of NMP. The washing step was repeated thrice and finally resuspended with 500 μl of NMP. A mass of 1 mg of azidoacetic acid *N-*hydroxysuccinimide ester was added to each well. The plates were then re-sealed with a chemically resistant silicone mat and shaken for 3 hours at 1500 rpm using a BioShake iQ thermal mixer. After azide modification, the previous wash steps were repeated, and the microparticles were finally resuspended in 500 μl of NMP. A mass of 0.5 mg of DBCO-PEG5-tetrafluorophenyl ester was added to each well, re-sealed with a chemically resistant silicone mat, and shaken for 5 hours at 1500 rpm using a BioShake iQ thermal mixer. The plate was washed with NMP thrice and then resuspended with 200 μl of NMP.

Barcode combinations were dispensed in a 96-well plate using an Echo 550 liquid handler and then transferred to the 96-deep-well plate containing the encapsulated microparticles. To each well of the 96 deep-well plates was added 800 μl of 50 mM carbonate buffer. The plate was re-sealed with a chemically resistant silicone mat and then shaken for 12 hours at 1500 rpm using a BioShake iQ thermal mixer. The barcoded microparticles were centrifuged at 1000 rcf then the supernatant was removed. The resulting pellet was washed with 20 mM Tris, 1 mM EDTA, and 0.1% (v/v) Tween 20 through repeated centrifugation, removal of the supernatant, and redispersing of the supernatant with 20 mM Tris, 1 mM EDTA, and 0.1% Tween 20 for three times. The microparticles were finally redispersed in 1000 μl of 20 mM Tris, 1 mM EDTA, and 0.1% (v/v) Tween 20. A volume of 500 μl from each well was taken and pooled together to create the sample library. No barcode degradation was observed over 4 months of room-temperature storage.

### Selection of microparticles

An aliquot of 500 μl of the sample library was placed in a 1.5-ml tube and then centrifuged at 1000 rcf to sediment the microparticles. The supernatant was removed then the microparticles were re-dispersed with 200 μl of hybridization buffer (10× SSC, 10% (v/v) dextran sulfate, 10% (v/v) formamide, and 0.05% (v/v) Tween 20). Separately, equivolume of barcodes probes, adapters, and fluorescent probes, and 1× SSC were pre-hybridized using the following method: 98°C for 10 seconds, 40°C for 5 minutes, 20°C for 2 minutes. A volume of 20 μl of prehybridized probe solutions were then added to the microparticle suspension. The resulting mixture was shaken at 1500 rpm for 15 minutes at 35°C using a BioShake iQ thermal mixer then centrifuged at 1000 rcf to sediment the microparticles. The supernatant was removed, and the microparticles were re-dispersed 1000 μl with the sorting buffer, composed of 1× SSC and 0.05% (v/v) Tween 20, to wash the microparticles. The microparticle sedimentation and washing steps were repeated thrice. The microparticles were finally resuspended in 1000 μl of sorting buffer. The fluorescently labeled microparticles were sorted using a Sony SH800 sorter equipped with a 100-μm sorting chip.

In the multi-pass selection process, existing fluorescent probes from previously sorted populations were meticulously removed before initiating subsequent rounds. Populations from an earlier selection were initially centrifuged in a 1.5-ml tube at 1000 rcf for 20 seconds, after which the sheath buffer was gently discarded. Next, 1000 µl of a denaturation buffer, comprised of 0.2 M NaOH in 90% formamide and pre-heated to 70°C, was added. This mixture was swiftly vortexed for 5 seconds and then incubated at 70°C in a BioShake iQ thermal mixer for 15 minutes. Following this incubation period, another 20-second centrifugation at 1000 rcf was performed then denaturation buffer was discarded. Then, 1000 µl of a denaturation wash buffer, composed of 0.05% Tween 20 and pre-heated to 70°C, was added. After a brief 5-second vortex and a 20-second centrifugation at 1000 rcf, the denaturation wash buffer supernatant was carefully siphoned off. Finally, 200 µl of hybridization buffer was added, preparing the sample for the next selection phase.

Sorted microparticles were de-encapsulated using 10 μl of electronics-grade 5:1 buffered oxide etch (VWR, catalog number: JT5192-3) and then diluted to 50 μl with nuclease-free water. The released samples were immediately used for Illumina sequencing library preparation.

### Internal barcode validation using short-read sequencing

A volume of 1 μl of 50 μM of combined master forward (5’– TCGTCGGCAGCGTCAGATGTGTATAAGAGACAGNNNNNGGCTATGAGACTGTTCGCTAAT* C*A*C –3’) and reverse (5’– GTCTCGTGGGCTCGGAGATGTGTATAAGAGACAGGAGACTAAACTGTGCCCACAAA*G*G* G –3’) primers with three consecutive phosphorothioates from the 3’-end, 44 μl of nuclease-free water, and 50 μl of repliQa HiFi ToughMix (Quantabio, catalog number: 95200-500) were added to 5 μl of released sample. The samples were amplified for 20 cycles using the manufacturer’s protocol then purified using 1× of AMPure XP beads (Beckman Coulter, catalog number: A63881). Samples were eluted from the magnetics beads using 22 μl of 20 mM Tris with 0.05% (v/v) Tween 20. Concentration of the amplified sample was measured using Qubit fluorescence assay (Thermo Fisher, catalog number: Q33231). Ten nanograms from the first PCR amplification were then taken to cycle-limited indexing PCR. Indexing PCR includes 10 μl of an indexing primer set from IDT® for Illumina® DNA/RNA UD Indexes (Illumina, catalog number: 20027213) or Nextera™ DNA CD Indexes (Illumina, catalog number: 20018708) and repliQa HiFi ToughMix as PCR master mix. Indexed samples were cleaned using 1× of AMPure XP beads and then quantified using quantitative PCR (qPCR). Final pooled libraries were then sequenced using an Illumina Nexsteq 2000 (800 pM loading concentration) using P1 flow cell with 150×2 reads, 50– 80% human genome spike-in for nucleotide diversity, and 2% PhiX internal standard.

### Synthetic SARS-CoV-2 sequencing

A portion of the released samples were then processed for sequencing SARS-CoV-2 samples using NEBNext® ARTIC SARS-CoV-2 FS Library Prep Kit for Illumina (New England Biolabs, catalog number: E7658) using the VarSkip primers with several modifications. Complementary DNA synthesis was performed using SuperScript^TM^ IV (catalog number: 18091200), 12 μl of released sample, and 1 μl of 50 μM 20-mer oligodeoxythymidine and 50 μM random hexamers for primers. Reverse transcription reactions were incubated at 50 °C for 1 hour. Finally, amplicons were amplified for 40 cycles.

Resulting libraries following the NEBNext® ARTIC SARS-CoV-2 FS Library Prep protocol were quantified using qPCR and then sequenced on a Nextseq 2000 (750 pM loading concentration) using a P1 flow cell with 150×2 reads and 1–10% PhiX internal standard.

Sequencing reads were first aligned with the SARS-CoV-2 Wuhan sequence (NC_045512.2) using minimap2^52^ (2.24-r1122). Human-readable sequence alignment maps were converted to binary alignment maps using samtools^53^ (v1.13). Resulting binary alignment map files were then used to demix the SARS-CoV-2 variants for each samples using Freyja^42^ (v1.4.5).

### Encapsulation of clinical SARS-CoV-2 samples

Copy numbers of the SARS-CoV-2 virus were measured upon receipt using probe-based quantitative reverse transcription polymer chain reaction (New England Biolabs; catalog number: M3019; Thermo Fisher; catalog number: A45583), detecting the N1 gene (Integrated DNA Technologies; catalog number: 10006713), and using synthetic SARS-CoV-2 Alpha variant as positive controls for the calibration curve (Twist Bioscience; catalog number: 103907). A sample volume of 1 µL was used for each qRT-PCR reaction.

To encapsulate, 500 µL of each sample was added to individual 1.5-mL tubes then diluted to 1000 µL using nuclease-free water. A mass of 1 mg of trimethylammonium-functionalized silica microparticles was then added to the solution. After mixing for 5 seconds using a vortex mixer, 10 µL of 50% *N*-[3-(trimethoxysilyl)propyl]-*N*,*N*,*N*-trimethylammonium chloride in methanol (TMAPS; Alfa Aesar; catalog number: H66414) and 10 µL of tetraethoxysilane (Millipore Sigma; catalog number: 333859) were then added. The resulting mixture was shaken using a BioShake iQ thermal mixer at 1600 rpm for 4 days, then centrifuged at 1000 rcf for 30 seconds to pellet the encapsulated samples. The supernatant was carefully removed, then 1000 µL of 2% (v/v) 2-azido-*N*-[3-(triethoxysilyl)propyl]acetamide in ethanol was added. The resulting mixture was further mixed for 16 hours at room temperature.

The azido-modified encapsulated samples were pelleted and washed twice with *N*,*N*-dimethylacetamide (DMAc; Millipore Sigma; catalog number: 185884) then the particles were re-dispersed with 1000 µL DMAc. A mass of 0.5 mg of DBCO-dPEG®₁₂-tetrafluorophenyl ester (Quanta Biodesign; catalog number: 11366) were added to each azide-modified encapsulated sample then the resulting mixture was shaken at 1600 rpm using a BioShake iQ thermal mixer for 1 hour at 40°C.

The tetrafluorophenyl-modified encapsulated samples were pelleted using a centrifuge at 1000 rcf for 30 seconds and washed twice with DMAc then the particles were re-dispersed with 100 µL DMAc. A volume of 900 µL of 500 mM phosphate buffer (Thermo Fisher; catalog number: J60825.AP) was added. Then, a volume of 10 µL of each DNA barcode as amino-modified DNA oligonucleotides 500 µM in nuclease-free water were added to each sample. Three barcodes for each sample were used to add complexity to the library. The table below shows the DNA barcode assigned to each sample.

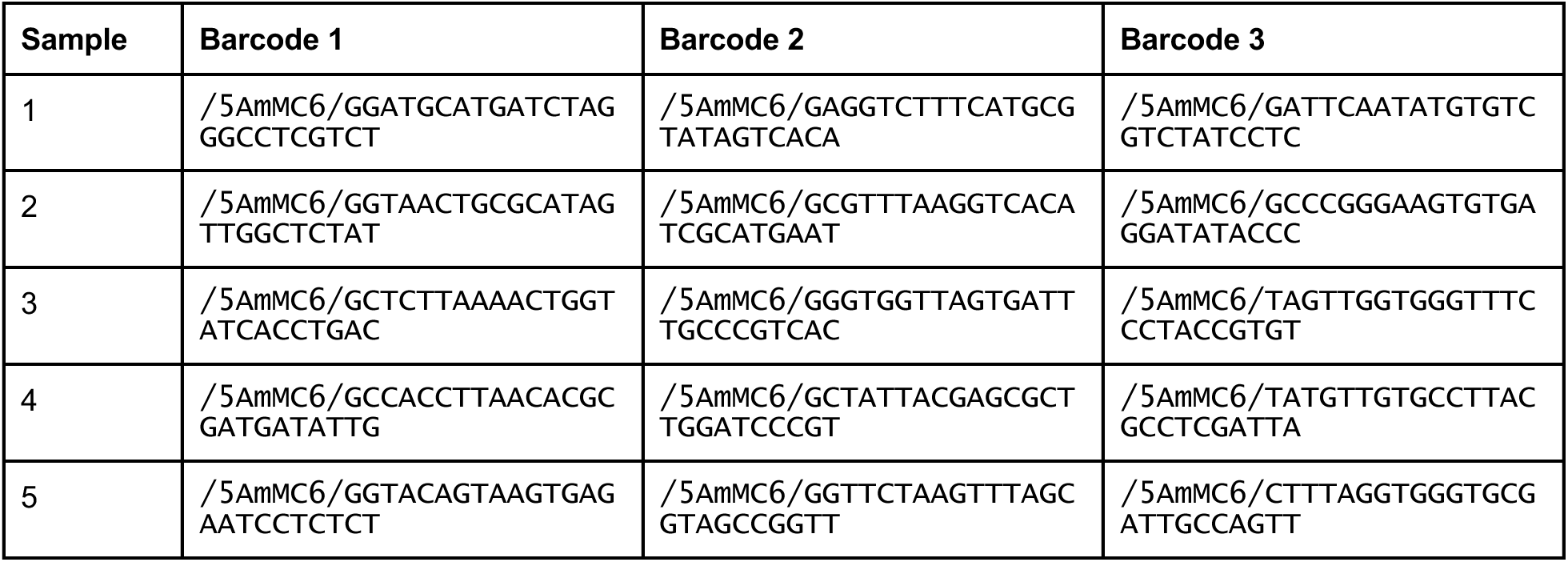

After 16 hours of mixing on a thermomixer at room temperature, the barcoded encapsulated samples were pelleted using a centrifuge at 1000 rcf for 30 seconds, washed twice with 1000 µL of hybridization buffer, then finally resuspended with 1000 µl of hybridization buffer. Barcoded encapsulated samples were kept at room temperature.

### Clinical SARS-CoV-2 sequencing

Ten µl of unencapsulated and 500,000 microparticles of encapsulated clinical SARS-CoV-2 samples were prepared for sequencing using NEBNext® ARTIC SARS-CoV-2 FS Library Prep Kit for Illumina (New England Biolabs, catalog number: E7658) using the VarSkip primers with several modifications. Encapsulated RNA was released from microparticles with 25 µl of 5:1 buffered oxide etch, and 12 µl was desalted using a 7k MWCO Zeba column (Thermo Fisher; catalog number: 89878). To remove any residual DNA fragments, all samples were first subjected to DNAse treatment (Thermo Fisher; catalog number: 11766051). First-strand complementary DNA synthesis was performed using SuperScript IV, 12 µL of the released sample or unencapsulated sample, and 20-mer oligodeoxythymidine and random for primers. Reverse transcription reactions were incubated at 50°C for 1 hour. Finally, amplicons were amplified for 40 cycles.

Following the rest of the NEBNext® ARTIC SARS-CoV-2 FS Library Prep protocol, the resulting libraries were quantified using qPCR and then sequenced on a Nextseq 2000 (800 pM loading concentration) using a P3 flow cell with 150×2 reads and 2–20% PhiX internal standard.

Sequencing reads were aligned using bwa^54^ (v.0.7.17-r1188). Sequence alignment maps were then converted to binary alignment maps using GATK^55^ (v4.3.0.0). Variant calling was performed using samtools^53^ (v1.13). Consensus sequences were generated using ivar^56^ (v1.3.1). Finally, SARS-CoV-2 lineages were analyzed using NextClade^57^ (v2.12.0).

For variant calling precision and recall analyses, duplicates from binary alignment maps were filtered using GATK MarkDuplicatesSpark then variant calling was performed using GATK HaplotypeCaller. Variants were filtered using GATK VariantFiltration. First, variants with a QualByDepth (QD) value less than 2.0 were excluded, using a filter tag “QD2”. QD provides a normalized variant confidence score by the depth of sample reads supporting a variant. Variants with a raw quality score (QUAL) below 30.0 were discarded, labeled under the “QUAL30” filter. Strand Odds Ratio (SOR), a metric that denotes the symmetry of the variant’s presence in both forward and reverse reads, was also considered. Variants with an SOR greater than 3.0 were filtered out and marked with the “SOR3” tag. This ensures that the variant is supported by both forward and reverse reads and isn’t an artifact from a potential strand bias. Further, Fisher Strand (FS) values, which indicate strand bias, exceeding 60.0 led to excluding the respective variants, tagged under the “FS60” filter. Lastly, variants with a Mapping Quality (MQ) less than 40.0, indicative of the overall alignment quality of reads supporting a given variant, were filtered out and designated with the “MQ40” tag.

Normalization of filtered variant calling files and variant overlap analyses between encapsulated and unencapsulated samples were performed using BCFtools^28^ (v1.13). True positive (TP) variant counts were directly inferred from the overlapping VCF, while the false positive (FP) and false negative (FN) counts were derived by subtracting TP from the encapsulated and unencapsulated normalized variant calling files, respectively. Precision was computed as the proportion of TP relative to the sum of TP and FP, and recall was derived as the proportion of TP relative to the sum of TP and FN.

## Data Availability

All data produced in the present study are available upon reasonable request to the authors.

https://doi.org/10.5281/zenodo.10501348

## ACKNOWLEDGEMENTS

M.B. and J.D.B. were supported by the Office of Naval Research (N00014-21-1-4013), the Army Research Office (ICB Subaward KK1954), and the National Science Foundation (CBET-1729397, OAC-1940231, and CCF-1956054). Additional funding to J.D.B. was provided through a National Science Foundation Graduate Research Fellowship (Grant No. 1122374). J.L.B. acknowledges support in part by the National Science Foundation SBIR Phase I 2136447, UCSF Parnassus Flow CoLab RRID:SCR_018206, DRC Center Grant NIH P30 DK063720, UCSF Center for Advanced Technology at Mission Bay, and Illumina. This research was also supported by a core center grant from the National Institute of Environmental Health Sciences, National Institutes of Health (P30-ES002109). We are grateful to T. B. Schardl and C. E. Leiserson for useful discussion about DNA barcoding. We thank G. Paradis, M. Jennings, and M. Griffin of the Flow Cytometry Core at the Koch Institute at the Massachusetts Institute of Technology (MIT) for flow sorting assistance. We are grateful to Delaware Diagnostics Labs for providing us de-identified clinical SARS-CoV-2 samples. We thank G. Tikhorimov for providing access to a Beckman Coulter Labcyte Echo 550.

## ETHICS DECLARATION STATEMENT

De-identified clinical samples were sourced from Delaware Diagnostic Labs. The MIT Committee on the Use of Humans as Experimental Subjects determined that IRB approval was not required for this study.

